# Coregulated metabolite networks associated with global protein crotonylation are central pathophysiological processes in prediabetes and diabetes

**DOI:** 10.64898/2026.04.19.26351178

**Authors:** Dhruv Dubey, Tumpa Dutta, Anna Casu, Anton Iliuk, Stephen J. Gardell, Richard E. Pratley, Yury O. Nunez Lopez

**Author notes:** Correspondence &. Brigham and Women’s Hospital/Harvard Medical School, Boston, MA 02115, USA.

## Abstract

Type 2 diabetes and prediabetes affect hundreds of millions of people globally, yet the metabolic networks underlying disease development remain poorly understood. Using untargeted liquid chromatography-mass spectrometry (LC-MS/MS), we profiled a total of 15,470 (900 known) serum metabolite features across the human diabetes spectrum (the most comprehensive coverage reported to date). Weighted coexpression network analysis of samples from people with normal glucose tolerance, prediabetes, and type 2 diabetes, collected at baseline and 2 hours after an oral glucose tolerance test, revealed tightly coregulated modules strongly associated with glycemic dysregulation, insulin resistance, and islet dysfunction. Notably, short-chain organic acids, particularly crotonic acid, emerged as hubs of the diabetes-associated networks, accumulating progressively with disease severity. Reanalysis of extracellular vesicle proteomics from the same cohort showed that 16.5% of circulating proteins were crotonylated, with 47.6% correlated with crotonic acid and other hub metabolites, establishing a metabolome-crotonylome axis as a novel mechanism in diabetes development.

## Introduction

Affecting hundreds of millions of people globally, type 2 diabetes mellitus (T2D) and prediabetes are metabolic disorders that have reached epidemic proportions worldwide [1, 2]. T2D is a common metabolic disorder characterized by pancreatic β-cell dysfunction and insulin resistance, resulting in hyperglycemia leading to an increased risk of complications ranging from neuropathy and retinopathy to cardiovascular disease [3–5]. Prediabetes is a precursor to T2D and is characterized by glycemic levels that exceed normal ranges yet remain below the diabetes threshold [6]. The global prevalence of prediabetes and type 2 diabetes continues to rise, creating a need for further studies to understand the pathogenesis of diabetes development and progression [7, 8].

In the past, metabolomics has been applied to the study of diabetes to uncover novel biomarkers, increase our understanding of diabetes pathophysiology, and develop better therapeutics and interventions [9]. Compared to genomics and proteomics, metabolomics has proven itself as a more reliable method of biomarker identification due to increased accuracy in identifying pathophysiological changes within cells and systems [10, 11]. Additionally, studies have shown that combining metabolomic data with clinical data and other omic technologies such as proteomics enhances the predictive performance and clinical utility [12] as well as the study of causal associations [13]. Previous metabolomics studies have consistently identified branched chain amino acids, aromatic amino acids such as phenylalanine, and metabolites involved in carbohydrate and lipid metabolism as central components of the metabolic signature associated with insulin resistance and diabetes pathogenesis [14–18]. Notably, a recent 2026 landmark metabolome-wide association study integrating 469 metabolites in 23,634 prospective participants from 10 prospective cohorts identified 235 circulating metabolites associated with incident T2D risk, including 67 previously unreported associations spanning glycine/serine, bile acid, carnitine, and lipid pathways [19]. However, this effort did not address the coregulated network architecture of the metabolome and its dynamic response to glucose. Complementary evidence from the IMI-DIRECT study (n=3,000) previously identified novel markers of glycemic deterioration, including 3,4-dihydroxybutyric acid and N-lactoyl amino acids, pointing to understudied metabolites beyond the canonical branched-chain amino acids and lipid biomarkers as contributors to diabetes progression [20]. In general, many of these prior studies have only detected small subsets of metabolites and analyzed a limited metabolome under fasted conditions. Our research extends beyond these studies by providing a global unbiased outlook of the human metabolome in response to an oral glucose tolerance test (OGTT) and comparing this to baseline conditions in participants with normal glucose tolerance, prediabetes and established diabetes, providing unique insight on metabolomic changes across the spectrum of dysglycemia. We specifically employed liquid chromatography-mass spectrometry (LC-MS/MS) technology to conduct global metabolomic profiling due to its high sensitivity, comprehensive coverage, and resolution [21]. This procedure has enabled us to gain a more comprehensive view of the human metabolome and extends beyond the limitations of prior studies, which have only analyzed a relatively small subset of metabolites. Additionally, we employed novel computational and statistical methods, integrating metabolomic coexpression network analysis, extracellular vesicle (EV) protein crotonylation analysis, differential expression analysis, and functional enrichment analysis to discover complex metabolic signatures across the diabetes spectrum and elicit mechanistic insight while assessing metabolite and protein network dynamics. Our analysis is differentiated from prior metabolomics studies using OGTT samples due to the application of coexpression network analysis, which takes advantage of the power-enhancing, naturally occurring multiple correlations in metabolomic data [22] and the implementation of a multiomic approach integrating the analysis of global EV proteomic data that serves as validation of key metabolomic findings.

## Results

### Study Population

In this metabolomics study, a well-balanced subset of patients was carefully selected from the ORIGINS (Uncovering the ORIGINS of Diabetes) study (ClinicalTrials.gov identifier: NCT02226640). Our group has previously documented the parent study cohort [23]. In total, 30 participants were chosen (n=10 in each cohort: NGT, Pre-T2D, T2D), and confounding effects of age, sex, and obesity were minimized in each cohort during selection. Serum samples were obtained from the subjects at baseline and after OGTT (120min), and LC-MS/MS-based untargeted metabolomics was performed on these serum samples from each of these subjects before and a 2-h during the OGTT. This same cohort was previously used to identify proteomic and phosphoproteomic signatures in circulating extracellular vesicles and dissect EV-mediated mechanisms of diabetes development and progression [24]. The clinical characteristics of this cohort are presented in Table 1.

**Table 1.** Tables should be placed in the main text near the first time they are cited.

|  | **Healthy** | **PreT2D** | **T2D** | **p** |
| --- | --- | --- | --- | --- |
| n | 10 | 10 | 10 |  |
| Sex = Male (%) | 5 (50.0) | 5 (50.0) | 5 (50.0) | 1.000 |
| Age (years) | 45.9 (10.5) | 48.0 (7.8) | 50.7 (11.0) | 0.560 |
| BMI (kg/m<sup>2</sup>) | 33.1 (6.5) | 34.1 (5.6) | 34.7 (5.2) | 0.825 |
| Weight Average (kg) | 89.7 (16.7) | 101.0 (15.6) | 97.8 (21.4) | 0.368 |
| Height Average (cm) | 165.4 (7.8) | 172.8 (11.1) | 167.5 (9.6) | 0.223 |
| Waist Circumference (cm) | 99.6 (16.4) | 110.8 (15.8) | 109.63 (16.2) | 0.252 |
| DEXA Lean Mass (g) | 52541.6<br>(8827.6) | 56993.2<br>(12429.7) | 54111.7<br>(12128.5) | 0.672 |
| DEXA Fat Mass (g) | 34737.9<br>(16103.2) | 41785.8<br>(15705.2) | 41624.5<br>(12800.2) | 0.494 |
| DEX Fat Percentage (%) | 38.3 (14.0) | 41.7 (12.2) | 43.0 (9.3) | 0.672 |
| HDL (mg/dL) | 55.9 (11.8) | 42.2 (8.9) | 48.7 (16.6) | 0.076 |
| LDL (mg/dL) | 113.7 (26.5) | 113.6 (46.7) | 102.3 (37.0) | 0.741 |
| Triglycerides (mg/dL) | 105.0 (60.4) | 159.7 (134.1) | 139.5 (53.4) | 0.404 |
| TSH (mIU/L) | 1.6 (0.7) | 2.1 (1.1) | 2.4 (1.1) | 0.180 |
| Temperature (F) | 98.0 (0.3) | 97.9 (0.4) | 97.8 (0.2) | 0.605 |
| Respiration Rate (breaths per min) | 14.7 (2.1) | 15.5 (1.8) | 15.2 (1.9) | 0.646 |
| Systolic Blood Pressure (mmHg) | 124.4 (10.1) | 125.2 (10.4) | 126.0 (11.9) | 0.950 |
| Diastolic Blood Pressure | 78.30 (9.1) | 77.2 (9.9) | 79.80 (8.2) | 0.815 |
| (mmHg) |  |  |  |  |
| Heart Rate (beats per min) | 63.4 (8.4) | 71.0 (11.5) | 67.6 (13.4) | 0.340 |
| HbA1C (%) | 5.4 [5.3, 5.5] | 5.9 [5.8, 6.1] | 6.3 [6.1, 7.0] | 0.001 |
| Glucose baseline (mg/dL) | 90.3 (6.7) | 100.6 (8.2) | 122.2 (20.3) | <0.001 |
| Insulin baseline (μIU/mL) | 3.6 (2.2) | 8.6 (6.8) | 5.8 (5.8) | 0.134 |
| Glucose AUC (mg/dL · min) | 14797.3<br>[13302.1, 16433.5] | 17202.3<br>[16488.6, 18909.1] | 25796.2<br>[23196.5, 29180.5] | <0.001 |
| Insulin AUC (μIU/mL · min) | 3594.0 [2560.8, 7438.8] | 6271.0 [2964.6, 11626.3] | 3463.6 [2254.2, 4915.9] | 0.237 |
| C-peptide AUC (ng/mL · min) | 694.2 [569.6, 820.0] | 950.3 [573.5, 1262.8] | 816.4 [687.6, 887.7] | 0.783 |
| Insulinogenic Index (ΔIns<sub>0-30min</sub>/ΔGlu<sub>0-30min</sub>) | 0.9 (0.6) | 0.3 (3.0) | 0.2 (0.2) | 0.623 |
| HOMA IR | 0.8 (0.5) | 2.1 (1.5) | 2.0 (2.2) | 0.155 |
| HOMA_B | 51.5 (36.4) | 96.0 (107.6) | 30.7 (22.5) | 0.102 |
| MATSUDA | 12.2 (9.4) | 6.3 (5.5) | 9.7 (8.9) | 0.276 |
| AIRg | 468.6 (349.8) | 549.32 (530.1) | 48.75 (64.6) | 0.041 |
| Si | 4.6 (4.5) | 2.7 (2.9) | 4.6 (2.6) | 0.434 |
| DI | 1150.7 (494.2) | 890.3 (569.4) | 249.5 (348.3) | 0.005 |

*Data presented as Mean (Standard Deviation) or Median [Interquartile Range]. Sex as number of males (%)*

### Global Serum Metabolomics of the Diabetes Spectrum

By conducting LC-MS/MS untargeted metabolomic profiling on the serum samples, we detected a total of 15,470 metabolite features, known and unknown, across all samples. Multiple data acquisition modes were used to detect these metabolites, including the positive and negative modes of HILIC and reverse phase C18. The expression of metabolites detected in more than one mode was averaged. Of the 15,470 metabolite features, 900 were known and 709 were known and detected in more than 70% of the samples. This subset of 709 known metabolites was used in subsequent analyses to identify differentially expressed metabolites and construct coexpression networks.

### Coexpressed Metabolite Networks Associated with Diabetes Traits at Baseline and in Response to OGTT

To fill a gap dissecting the complex metabolic networks at play during diabetes development, we conducted an analysis of metabolite coexpression networks using WGCNA [25]. Using the WGCNA wrapper *BioNERO* [26], key metabolite modules were identified at baseline and 2 hours after an oral glucose tolerance test. In particular, we focused on a “Baseline” network that mapped metabolite associations at time zero of an oral glucose tolerance test and a “Delta” network that mapped the metabolite changes from baseline to 2-hours during the glucose tolerance tests (Figure 1). We assigned particular importance to network modules that correlate with clinical and physiological traits relevant to the progression of diabetes (Figure 2).

**Figure 1.**
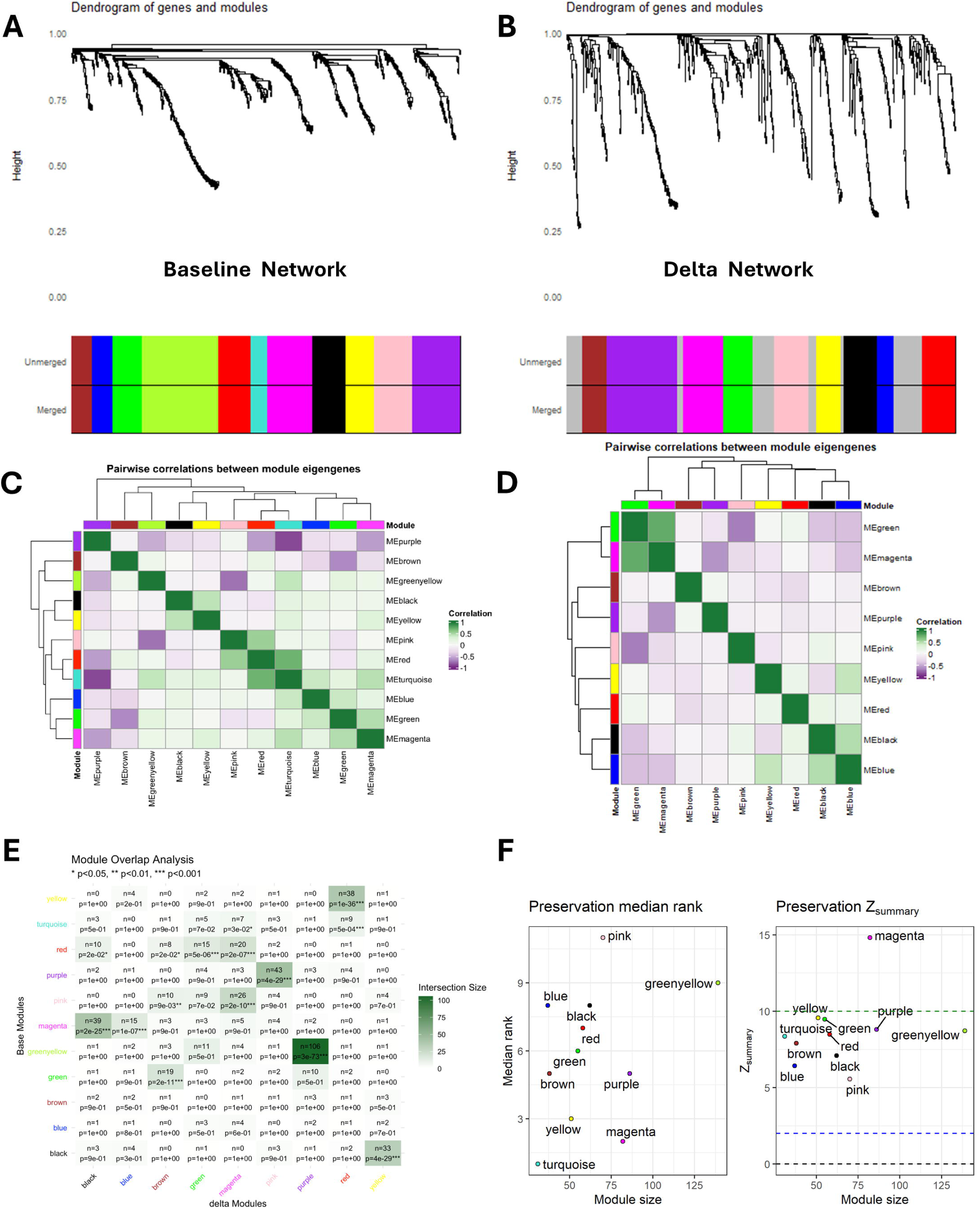
Weighted metabolite coexpression networks. Clustering dendrograms for Baseline (A) and Delta (B) networks. The y-axis represents a dissimilarity distance (1 - TO), and the x-axis depicts modules with their colors as determined by dynamic tree cutting at a predetermined height. Pairwise correlations between module eigengenes in Baseline (C) and Delta (D) networks. The module eigengene describes a representative metabolite expression profile of a module. A stronger pairwise correlation between two modules indicates greater module similarity. Correspondence between Baseline and Delta networks (E): Crosstabulation of metabolite identities between all pairwise module comparisons. Overlap between modules is reported as the number of common metabolites. P-values are represented by *** <0.001, ** <0.01, and * < 0.05. Preservation analysis of baseline network modules (F).

**Figure 2.**
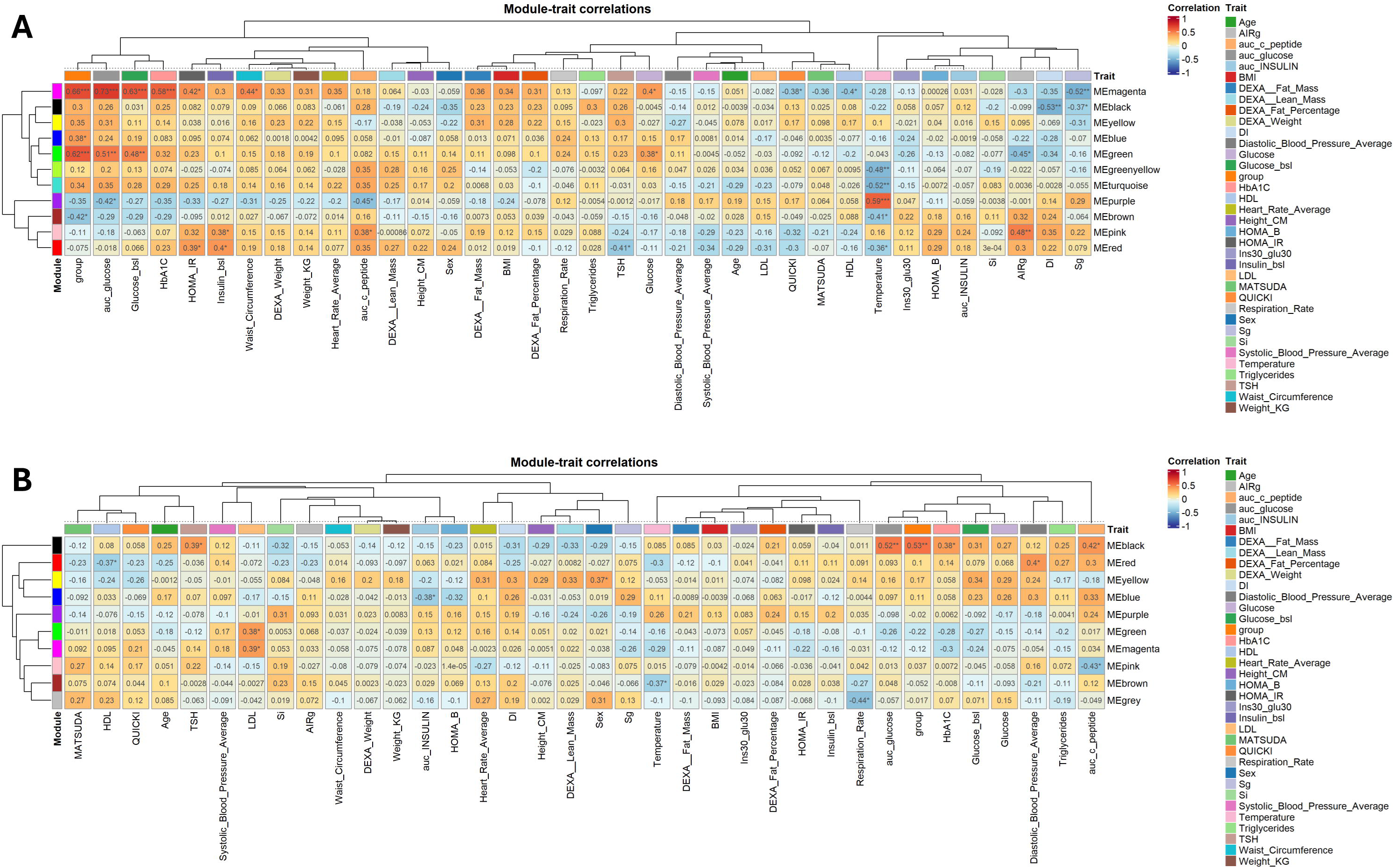
Module - Trait correlations. The identified modules in Baseline (A) and Delta (B) networks were correlated with clinical variables. Correlation coefficients are reported. P-values represented by *** <0.001, ** <0.01, and * < 0.05.

Specifically, we identified 11 modules in the Baseline network and 9 modules in the Delta network (Figure 1). Some of these modules, like the Baseline Greenyellow and the Delta Purple modules, and the Baseline Black and the Delta Yellow modules, overlapped between the two networks (see the cross-tabulation of modules metabolites to infer correspondence between the Baseline and Delta modules, Figure 1E).

Of note, eight of the Baseline modules (black, blue, brown, green, magenta, pink, purple, red) were significantly (P < 0.05) correlated with at least one of the following diabetes-related traits: diabetes group, fasting glucose, area under curve for glucose (AUC-Glucose), HbA1c, HOMA IR, fasting insulin, disposition index, and glucose effectiveness (Figure 2). On the other hand, the Delta Black module correlated with several glycemic control traits [i.e., AUC-Glucose (r=0.52, P<0.01), HbA1c (r=0.38, P<0.05), and the Diabetes group (r=0.53, P<0.01)], while the Delta Blue module negatively correlated with AUC-insulin (r=-0.38, P<0.05). The Delta Black and Delta Blue modules segregated with the key Baseline Magenta module (see cross-tabulation of modules metabolites in Figure 1E). Both the Delta Black and Delta Blue modules showed substantial overlap (39 metabolites, p = 2x10^-25^ and 15 metabolites, p = 1x10^-7^, respectively) with the Baseline Magenta module and were significantly correlated with relevant clinical measures. This suggests that the coregulated metabolites from the Baseline Magenta module segregate into two main global pathways in response to the oral glucose challenge, with relevance to diabetes pathophysiology. Also notable is the fact that short/medium-chain organic acids are key hub metabolites in these modules associated with the diabetes trait and measures of glycemic control and insulin secretion and action. Among those short-chain organic acids were aldehyde acids (e.g, glyoxylic acid), ketoacids (e.g, acetoacetic acid), and short-chain fatty acids (SCFA: e.g, crotonic acid and 3S,4-dihydroxy-butyric acid), among others (Figure 3).

**Figure 3.**
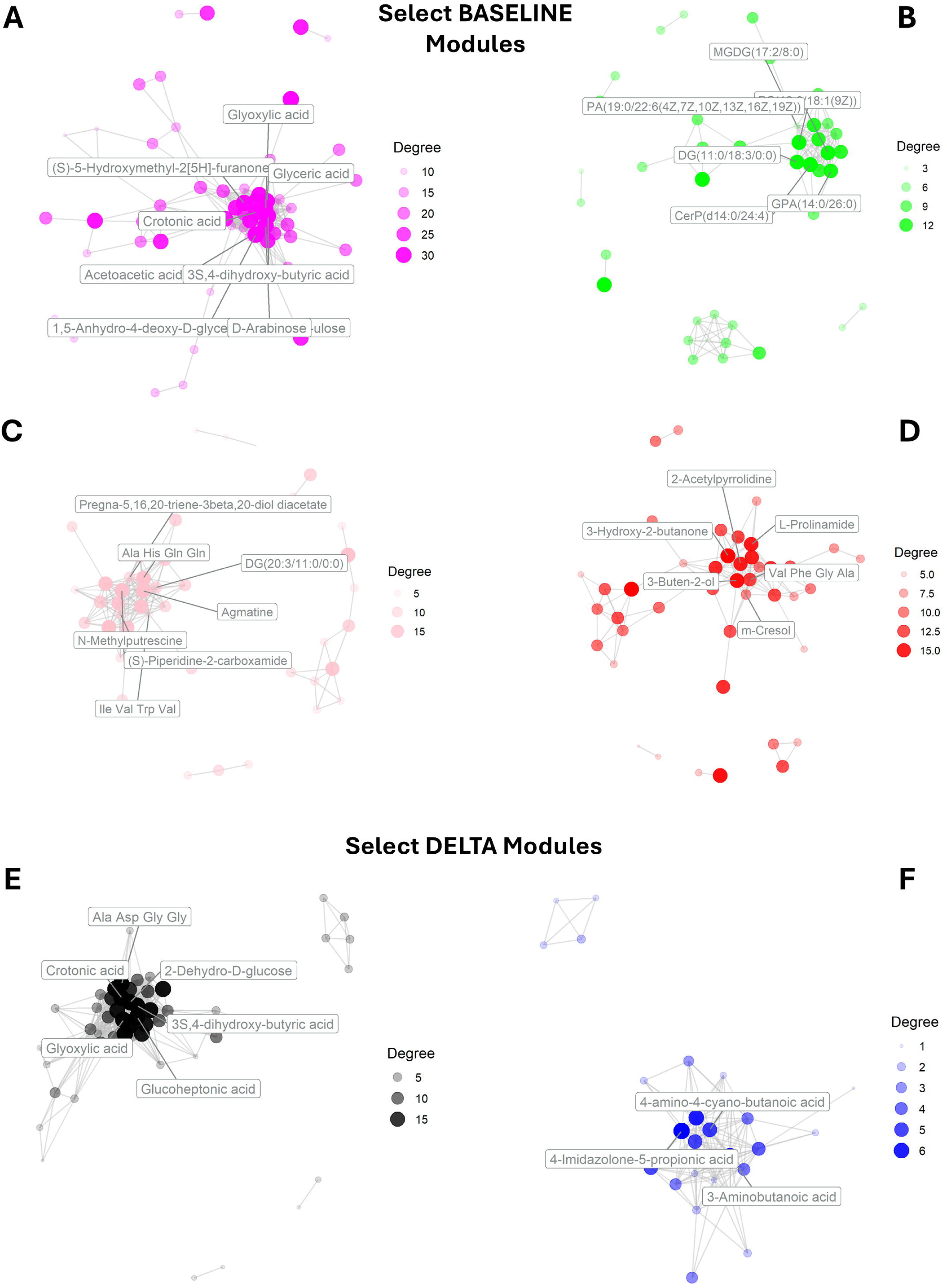
Subnetworks of significant (P<0.05) intramodular correlations in select modules associated with measures of glycemic control, pancreas function, and/or diabetes trait: Baseline Magenta (A), Baseline Green (B), Baseline Red (C), Baseline Pink (D), Delta Black (E), and Delta Blue (F). Hub metabolites are labeled on each network. kWithin: intramodular connectivity of a gene within a module (calculated as the sum of the connection strengths between a metabolite and all other metabolites within the same module), MM: module membership (range 0 to 1).

According to the module preservation analysis computed by BioNERO, the Baseline Magenta module (comprised of n=86 metabolites) is, overall, the top most preserved module from the Baseline network, as compared to the modules in the Delta network (Figure 1F). This means that the majority of correlations among Magenta module metabolites remains after the glucose challenge. Importantly, the Baseline Magenta module displayed a coexpression pattern that was highly significantly correlated with the diabetes trait (r=0.66 P=6.5x10^-5^), AUC-glucose (r=0.73 P<0.001), and negatively correlated with measures of glucose effectiveness (Sg, r=0.52, P<0.01) (Figure 2A), among other relevant variables.

Another notable Baseline module was the Green module (n=55), which also strongly correlated with the diabetes trait (r=0.62, P=2.4x10^-4^) and glycemic control measures. The moderate preservation score for this module likely indicates a network of diabetes-associated metabolites that undergo a more heterogeneous response to the OGTT challenge. In analyzing the metabolites in the Baseline Green (B), several lipid species, including various glycerophospholipids (GPA, GPCho, PA, PC, PG), diacylglycerols (DG), and ceramide phosphates (CerP) emerged as potentially relevant hubs (Figure 3B) in diabetes pathophysiology.

Notably, many of these lipid species contain polyunsaturated fatty acid chains of various chain lengths, suggesting potential involvement in cell membrane dynamics and lipid signaling pathways. Both the Baseline Magenta and Baseline Green modules showed dynamic shifts across their metabolite composition in response to the glucose challenge, affecting about half of the detected metabolites and segregating into multiple Delta modules (Figure 1E).

On the other hand, the Baseline Pink module positively correlated with beta cell function measures and represents the most divergent module in response to the glucose challenge (Figure 1F). This module is characterized by the presence of hub metabolites such as N-methylputrescine, agmatine, and piperidine 2-carboxamide (Figure 3C) that are protective against diabetes and appears to be involved in mitochondrial beta oxidation of very long chain fatty acids (VLCFAs) consistent with fatty acid metabolism. As shown in Figure 1C and 2A, the Baseline Pink and the Baseline Red modules appear to be related based on their correlation distance and to coalesce into the Delta Magenta module, which suggests coregulated changes in response to the glucose challenge. The Baseline Red module is also enriched in metabolites related to beta oxidation, in this case, of branched-chain fatty acids (BCFAs), and in metabolites of the carnitine synthesis pathway (Figure 3D).

### Distinct Metabolic Pathways were Enriched in Diabetes-related Modules

To characterize the biological mechanisms and pathways underlying the identified coregulated modules, we conducted metabolite set enrichment analysis (MSEA) using the MetaboAnalystR software [27]. This tool maps the metabolites from each module to their corresponding annotation on The Small Molecule Pathway Database (SMPDB)’s comprehensive repository of metabolic pathways, enabling us to assign functional context to these modules and to identify specific pathways that are perturbed or enriched in relation to the studied experimental conditions [28].

Enrichment analysis revealed several biological pathways and mechanisms related to the modules found in the Baseline and Delta networks, highlighting the dynamic metabolic response when faced with a glucose challenge (Figure 4, 5). The Baseline Magenta module was significantly enriched (P<0.05, FDR<0.2) for glycine and serine metabolism, the citric acid cycle, amino sugar metabolism, and the transfer of acetyl groups into mitochondria (Figure 4A).

**Figure 4.**
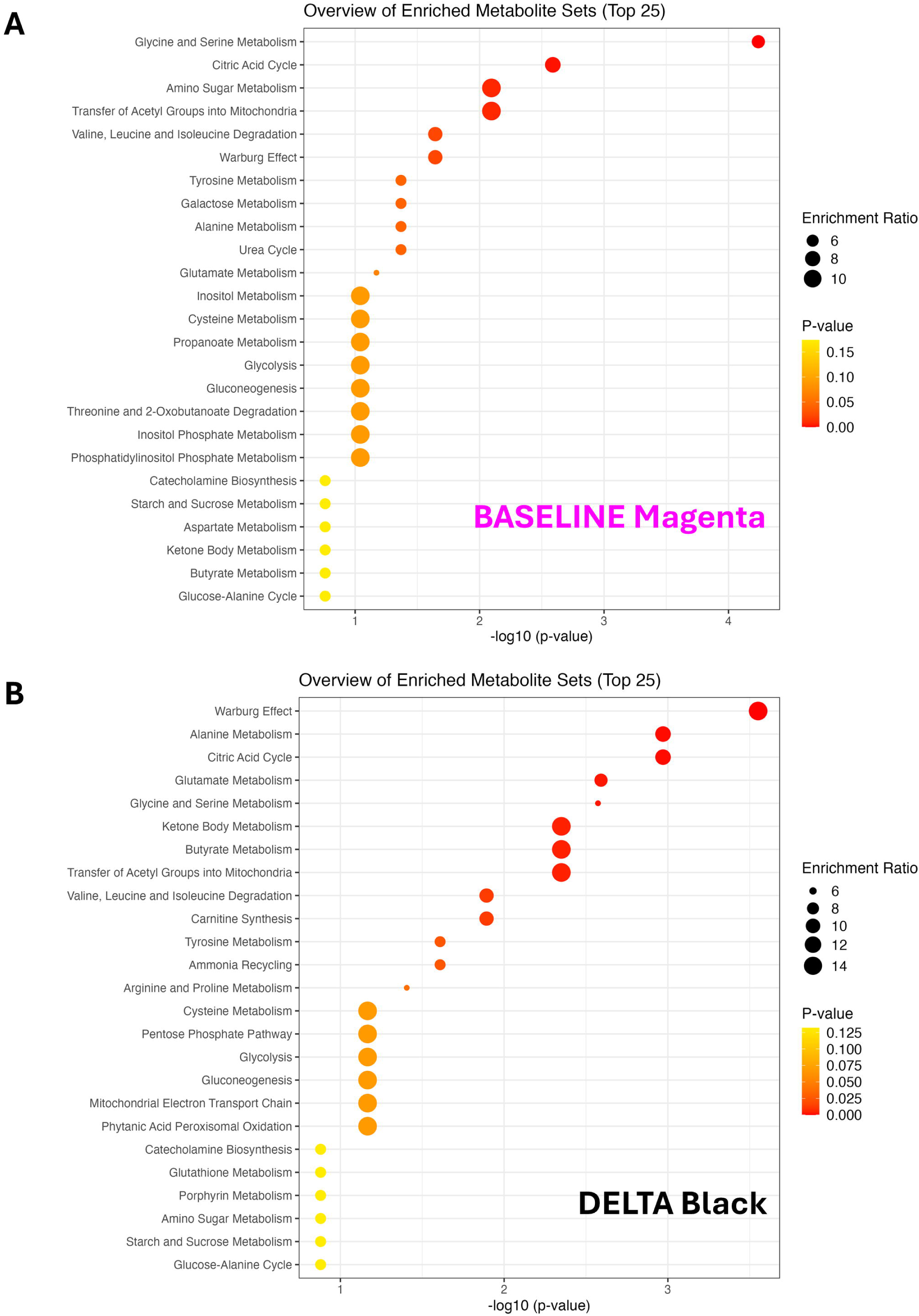
Enrichment analysis for the list of metabolites in the Baseline Magenta (A) and Delta Black (B) modules. Pathways collected from the Small Molecule Pathway Database (SMPDB). Metabolite set enrichment analysis (MSEA) was performed using the enrichment analysis R package MetaboAnalystR. The list of 709 identified metabolites was used as the background metabolite set.

**Figure 5.**
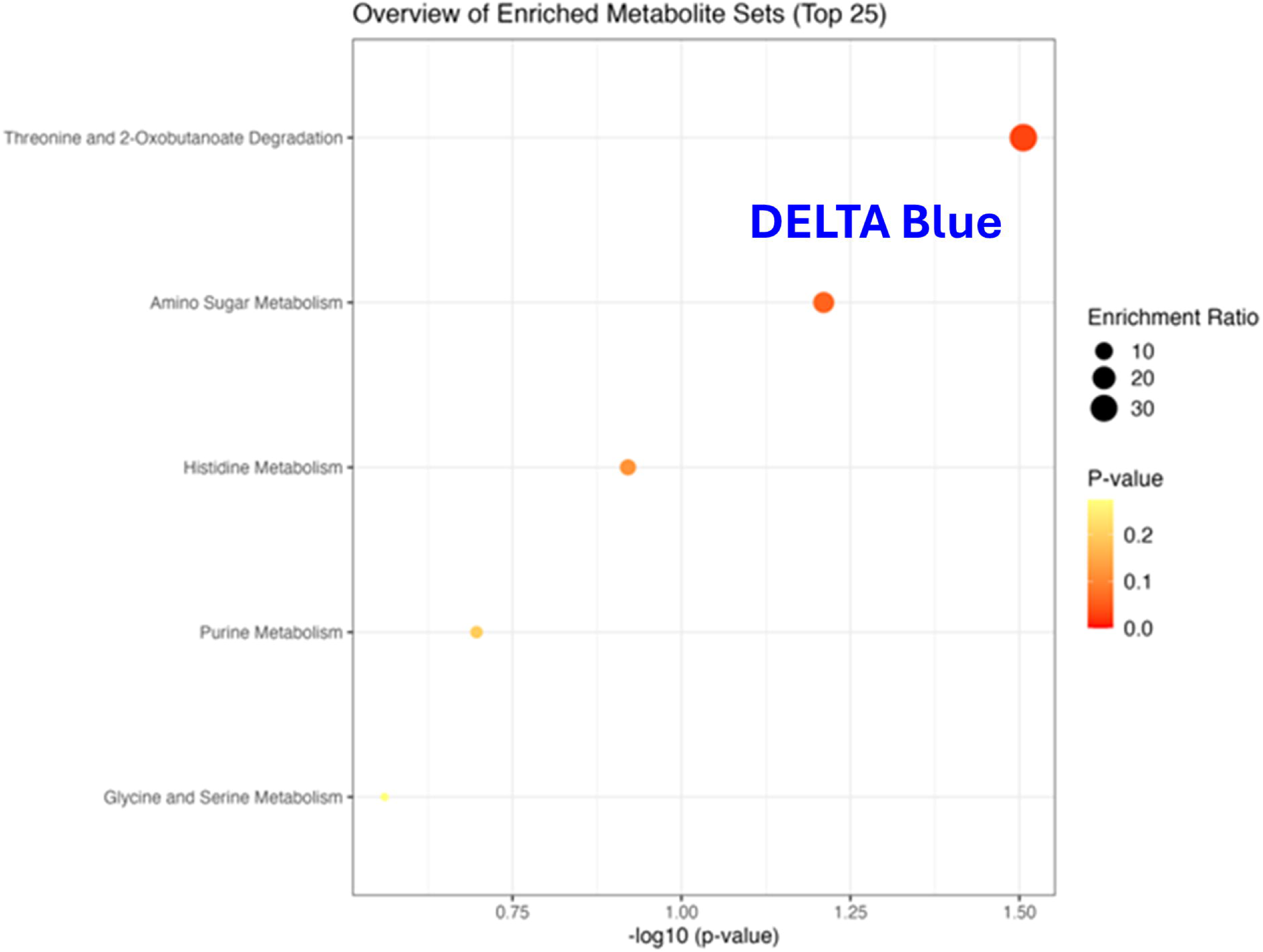
Enrichment analysis for the list of metabolites in the Delta Blue module. Pathways collected from the Small Molecule Pathway Database (SMPDB). Metabolite set enrichment analysis (MSEA) was performed using the enrichment analysis R package MetaboAnalystR. The list of 709 identified metabolites was used as the background metabolite set.

Showing a similar pattern to the Baseline Magenta module, the Delta Black module was most significantly enriched (P<0.05, FDR<0.2) not only for glycine and serine metabolism, the citric acid cycle, and the transfer of acetyl groups into mitochondria, but also for Warburg effect metabolites, ketone body metabolism, alanine and butyrate metabolism, degradation of BCAAs, and carnitine synthesis (Figure 4B).

### Differential abundance and interaction analysis of serum metabolites supports the progressive accumulation of organic acids and their derivatives in prediabetes and diabetes

To complement WGCNA and identify metabolite signatures that significantly changed (absolute fold change FC>1.5, P<0.05, FDR<0.05) among the study groups, we conducted differential abundance analysis using linear models that assessed both the main group effects at baseline and OGTT-120min, as well as the group-by-time (GxT) interaction effects (which test whether the molecular response to the glucose challenge differs across the glycemic spectrum), providing insight into the distinct physiological response to a glucose challenge in people with prediabetes and T2D, compared to normoglycemic individuals. Figures 6A and 6B and Supplementary Materials Tables S1-S9 display the signatures of all differentially abundant metabolites and metabolites with significant GxT interactions. Notably, all the hub metabolites identified in the

**Figure 6.**
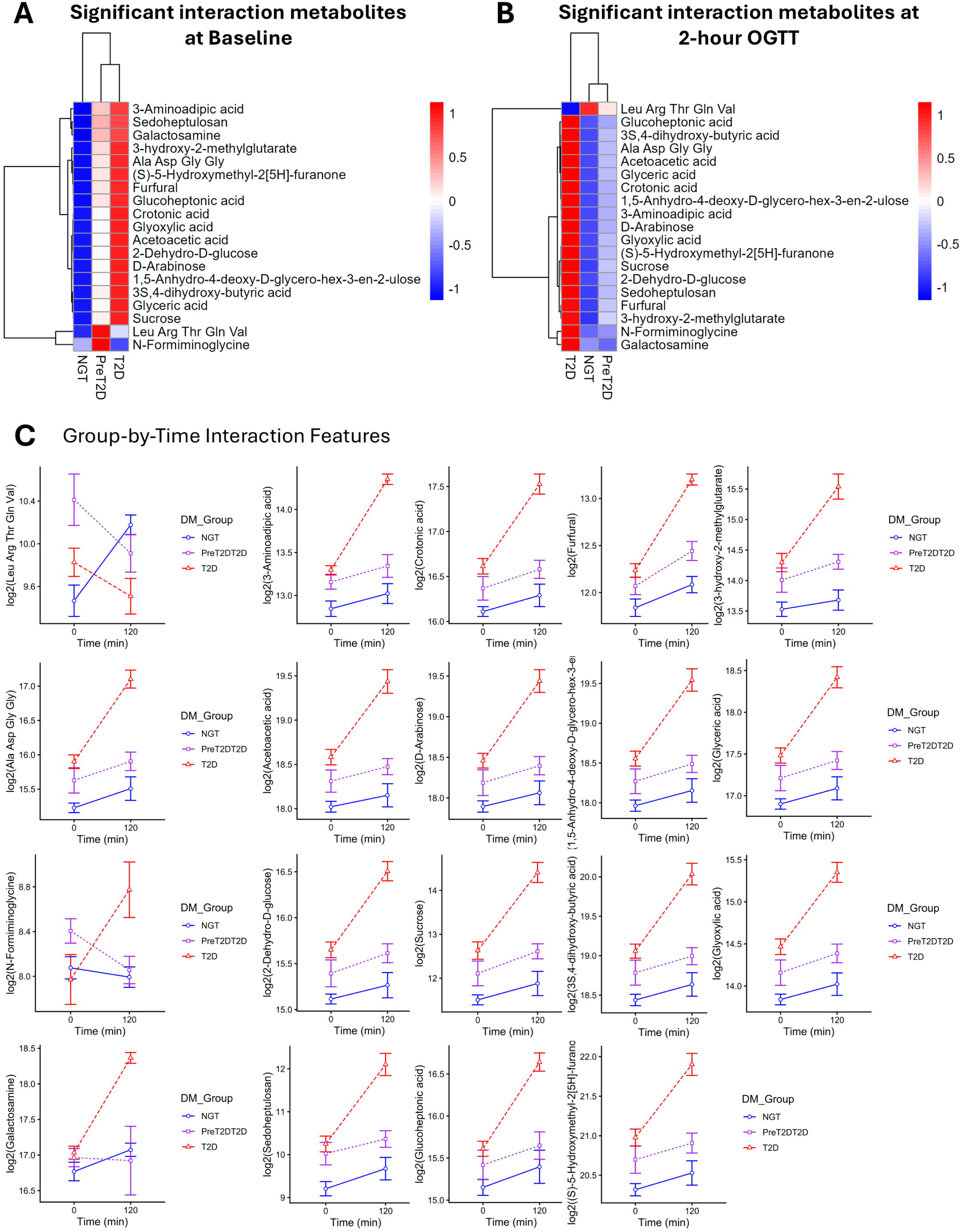
Differential expression analysis. Heatmaps of all metabolites with significant group-by-time interaction effects, plotted at baseline (A) and 120 minutes after an oral glucose tolerance test (OGTT) (B). Interaction plots of all metabolites with significant interaction effects (C). Analysis of differential metabolite abundance was conducted using linear models implemented with the *limma* package (*duplicateCorrelation* was calculated and used to model the random effects of the Subject_ID variable to account for the correlation between the repeated measures at baseline and OGTT-120min). Metabolomic features with fold change FC>1.5, P<0.05, and FDR<0.05 were considered significant.

Baseline Magenta module had significant GxT interaction. A majority of these Magenta module hub metabolites are short/medium-chain organic acids, including crotonic acid, glyoxylic acid, 3S,4-dihydroxy-butyric acid (DHBA), glyceric acid, and acetoacetic acid (Figure 3A and Figure 6C). Similarly, all but one of the hub metabolites in the Delta Black module belong to this group of metabolites with significant GxT interaction: crotonic acid, glyoxylic acid, 3S,4-dihydroxy-butyric acid, and glucoheptonic acid (Figure 3E and Figure 6C). The present overlap between these distinct analyses underscores the important role of these organic acids in diabetes.

Interestingly, one small peptide with the sequence Leu-Arg-Thr-Gln-Val was the only metabolite that displayed significant GxT interaction in the prediabetes group (compared to the response of the NGT group) under our stringent significance criteria (Figure 6C, Supplementary Materials Table S3).

Because DHBA is an intermediate in GABA metabolism, we also confirmed that GABA was significantly elevated in the T2D (although below our FC cutoff of 1.5, with FC=1.34, P=0.012, FDR=0.034), and that the metabolites display strong correlations at both baseline (r=0.45, P=0.013) and after the glucose challenge (r=0.4, P=0.03), as shown in Figure 7.

**Figure 7.**
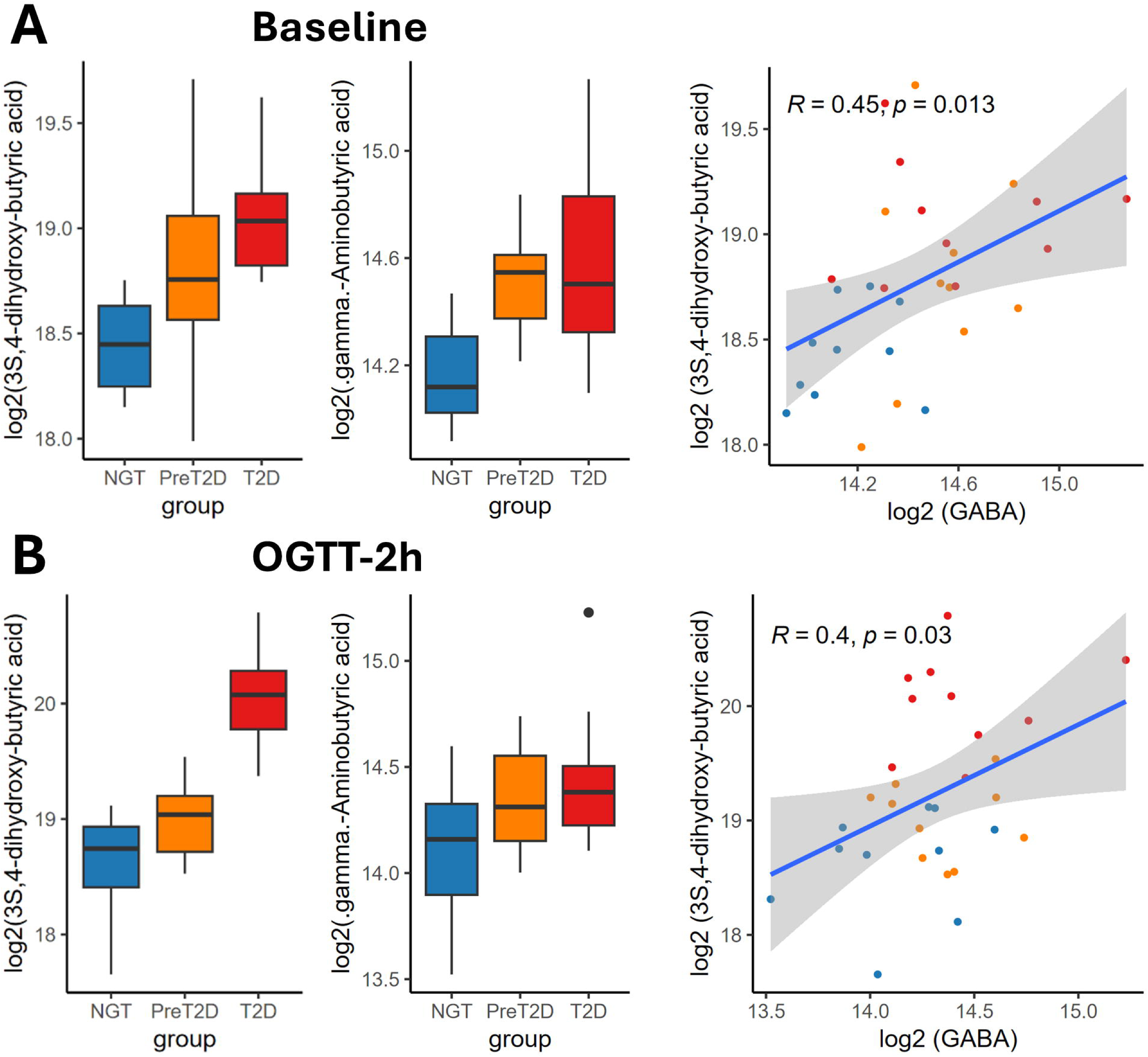
Expression of serum 3S,4-dihydroxy-butyric acid, GABA, and their respective correlation at baseline (A) and after the oral glucose challenge (OGTT) (B). Color-coded boxplots and scatter plots: NGT: blue, PreT2D: orange, T2D: red.

### Crotonylated proteins detected in serum extracellular vesicles significantly correlate with differentially abundant and hub metabolites including crotonic acid

Our results suggesting that crotonic acid plays a central role in diabetes, led us to reason that protein crotonylation should also be affected in diabetes and associated with the metabolomic signature. To validate this, we reanalyzed our previously published LC-MS/MS EV proteomic data [24] to identify the crotonyl post-translational modification in the EV proteins. Our rationale was that EVs represent, to some extent, a mirror image of their cells of origin, containing many of the proteins present in these cells in proportional concentrations, which are used for intercellular and inter-organ communication purposes. Other EV proteins destined for disposal may appear to be loaded in disproportionate concentrations. By quantifying crotonylated proteins in circulating EVs, we could gain insight into the systemic levels of crotonylated protein and their association with coregulated metabolite networks and clinical measurements.

Notably, we found that 397 serum EV proteins (16.5% of the detected global EV proteome) were crotonylated. Approximately two thirds (64% and 63%, respectively) of these EV crotonylated proteins tend to increase their levels in both prediabetes and T2D, as compared to the NGT group (Supplementary Materials Tables S10-S11). Because the ORIGINS study was not designed to assess global significance of crotonylated proteins, this impacted our statistical power to identify EV crotonylated proteins passing our stringent significant cutoffs after multiple testing correction for the group comparisons. However, we did found that 189 (roughly half the set) of those EV crotonylated proteins significantly (absolute /r/>0.4, P<0.05, FDR<0.1) correlated with the baseline levels of 166 network hub and significant differentially abundant metabolites (generating a total of 1,521 significant correlations, Supplementary Materials Table S12) that defined key global metabolic pathways associated with clinical measures of glucose control and beta cell function and that responded to the physiological glucose challenge. Serum levels of several crotonylated EV proteins significantly correlated with the serum levels of crotonic acid itself (Figure 8, Supplementary Material Table S13).

**Figure 8.**
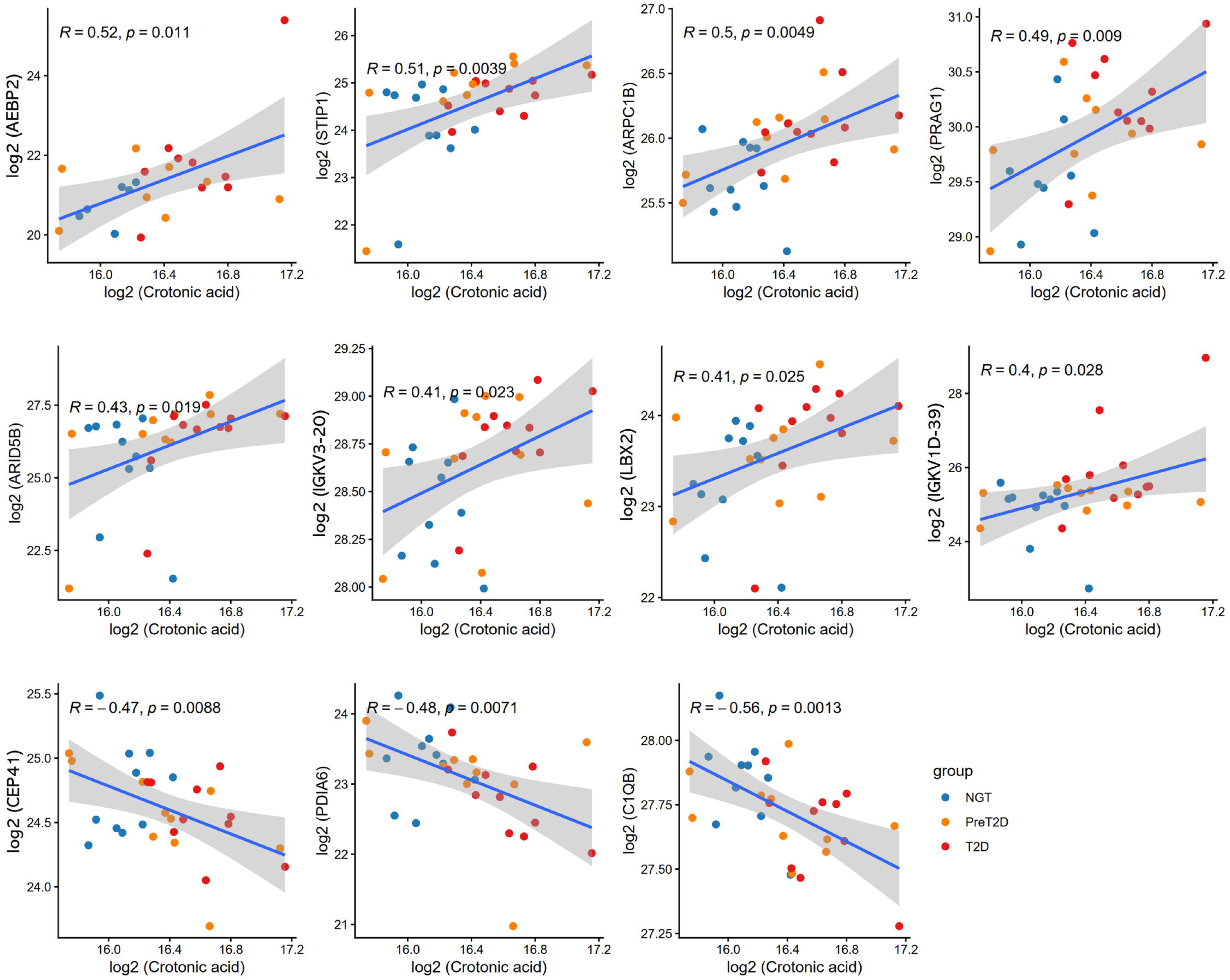
Significant correlations (absolute /r/>0.4, P<0.05, FDR<0.1) between baseline serum levels of crotonic acid and baseline serum levels of crotonylated EV cargo proteins. Color-coded scatter plots: NGT: blue, PreT2D: orange, T2D: red.

## Discussion

The present study provides one of the most comprehensive serum metabolomic characterization of the human diabetes spectrum to date, uncovering coregulated networks of metabolites that underlie the development of prediabetes and type 2 diabetes. A central and novel finding is that short-chain organic acid hubs, with a prominent role of crotonic acid, progressively accumulate with diabetes severity and appear to be mechanistically linked to global protein crotonylation in circulating EVs. This integrative multiomic approach, combining coexpression metabolomic network analysis with EV proteomics, reveals that the diabetic metabolome exerts a broad influence on the post-translational landscape of circulating EV proteins, offering a new mechanistic perspective on diabetes pathophysiology.

The coexpression network modules most significantly associated with diabetes correlated with glycemic control (fasting glucose, HbA1c, AUC-glucose), insulin resistance (HOMA-IR), and islet function (disposition index, AUC-insulin), and contained well-established diabetes metabolites such as branched-chain amino acids, aromatic amino acids (tyrosine, phenylalanine), glutamic acid, aminoadipic acid, ceramides, diacylglycerols, and bile acids among others, which provide cross-validation against the existing literature [14, 15, 29–35] and the recent 235 metabolites prospectively associated with incident T2D over 26 years of follow-up in 23,634 individuals [19]. Importantly, the Baseline Magenta module, the most strongly preserved module across both networks, segregated in response to the glucose challenge into two complementary Delta modules: the Delta Black (correlated with glycemic control) and Delta Blue (correlated with insulin secretion). This dissection suggests that the Baseline Magenta module encompasses two physiologically distinct subnetworks: one reflecting pancreatic endocrine function and one reflecting the composite metabolic response of the liver, muscle, and adipose tissue, that together constitute a central metabolic pathway of diabetes development.

A common feature of the Baseline Magenta and Delta Black modules is their enrichment in short-chain organic acid hubs that accumulate progressively in prediabetes and T2D, most prominently following the glucose challenge (particularly in the T2D group). This accumulation contrasts with a concurrent deregulation in the Baseline Pink and Red modules, which are enriched in mitochondrial beta-oxidation of very-long-chain and branched-chain fatty acids (VLCFAs/BCFAs), and are consistent with reports of impaired VLCFA catabolism in diabetes [36–39]. Together, these findings suggest a coordinated metabolic shift in which accumulating short-chain organic acids drive pathological network hubs while the long-chain fatty acid oxidation machinery becomes progressively impaired. In addition, and consistent with our identification of glycine and serine degradation as a prominently enriched pathway in the top diabetes-associated Baseline Magenta and Delta Black modules, a recent large-scale metabolome-wide association study independently identified glycine and L-serine metabolism as the top canonical pathway enriched among genetic loci of T2D-associated circulating metabolites, and further showed that circulating glycine mediates part of the protective association between coffee/tea consumption and incident T2D risk [19].

### Crotonic acid hub and a novel role for global protein crotonylation in diabetes

Among the most relevant short-chain organic acid hubs, identified by our study, was crotonic acid. Crotonic acid is a short-chain, unsaturated carboxylic acid that serves as the substrate for protein crotonylation, a post-translational modification first described in 2011, in which crotonyl groups are added to lysine residues [40]. Like butyrate, crotonic acid is likely derived from gut microbiota and once transported into cells via monocarboxylate transporters, it is converted to crotonyl-CoA for protein modification [41, 42]. Although crotonylation has been linked to kidney disease, neurological disorders, and cancers [41], its role in diabetes had not been established. Our study reveals that crotonic acid is a differentially abundant and progressively accumulating hub metabolite in the diabetes-associated network modules. To assess downstream effects, we identified 397 (16.5%) crotonylated serum EV proteins, of which 47.6% significantly correlated with hub metabolites including crotonic acid. These correlations are remarkable given the signal dilution expected in circulation, where many cell types contribute metabolites and EVs. Recent mechanistic studies provide compelling support. For example, Liu and colleagues showed that increased global protein crotonylation promotes white fat browning and activates mitochondrial energy metabolism proteins (AK2, Tpi1, NDUFA8) [43], potentially explaining increased resting energy expenditure reported in insulin-resistant conditions and in people with poor beta cell function [44, 45]. In addition, Liao and Zhang demonstrated that EPB41L4A-AS1, a long non-coding RNA elevated in T2D liver and insulin-resistant muscle, inhibits GLUT4 expression by reducing H3K27 crotonylation at the GLUT4 promoter [46]; and Shan et al. showed that *Enteromorpha prolifera* oligosaccharide (EPO) improves glucose metabolism by inhibiting HSPA8 crotonylation and activating the AKT pathway, while XPO1 crotonylation promotes cellular oxidative damage and aging [47]. Together, these findings establish elevated crotonic acid and consequent global protein crotonylation as a central, previously underappreciated mechanism in diabetes development and/or progression.

### Glyoxylic acid (glyoxylate)

Glyoxylic acid was a differentially abundant hub in both the Baseline Magenta and Delta Black modules. Produced via peroxisomal oxidation of glycolate or mitochondrial catabolism of hydroxyproline [48, 49], glyoxylate has emerged as a metabolic biomarker of T2D risk, with increased circulating levels detected up to 3 years prior to diagnosis [50]. In addition, elevated levels of glyoxylate are hypothesized to link excess fatty acid-to-glucose conversion to insulin resistance [50, 51]. Its co-elevation with arachidonic and dihomo-gamma linolenic acids in antihypertensive-treated T2D patients has been attributed to suppressed alanine:glyoxylate aminotransferase (AGT1/2) activity, linking glyoxylate to the intersection of diabetes and hypertension [52]. Our network-level identification of glyoxylate as a diabetes-associated hub reinforces its candidacy as an early biomarker and mediator of metabolic dysregulation warranting further validation.

### 3S,4-dihydroxy-butyric acid (3,4-DHBA)

3,4-DHBA, an intermediate in GABA metabolism and a hub in both the Baseline Magenta and Delta Black modules, was among the most significantly differentially abundant metabolites in our cohort. Given that GABA is produced in high concentrations by pancreatic beta cells [53], and that we confirmed elevated GABA in T2D (FC=1.34, P=0.012, FDR=0.034), the correlated accumulation of 3,4-DHBA and GABA in circulation suggests impaired pancreatic GABA clearance likely associated with disruption of mitochondrial oxidative function [54]. Recent independent validation for 3,4-DHBA as a biomarker of glycemic deterioration is provided by its significant association with HbA1c progression rate in the IMI-DIRECT cohort (n=3,000; β=0.060, FDR=0.00864) [20], confirming that this metabolite’s relevance extends beyond our cohort and across distinct study populations and metabolomic platforms. Additionally supporting its clinical relevance, 3,4-DHBA levels correlate with the stage of diabetic retinopathy and predict its longitudinal progression [55]. A Mendelian randomization study further implicated Christensenellaceae gut bacteria in elevated circulating 3,4-DHBA levels [56], suggesting this metabolite may bridge gut dysbiosis, diabetes, and its complications. Furthermore, the association of GABA with incident T2D risk [Hazard Ratio=1.07 (95%CI: 1.01-1.13), P=0.013, FDR=0.022] was also recently identified in a prospective metabolome-wide association study of 23,634 participants [19], which combined with our finding of co-elevated circulating GABA and 3,4-DHBA in prediabetes and T2D, adds support for the relevance of pancreatic GABA dysregulation as a feature of the diabetic state detectable in circulation.

### 4-imidazolone-5-propionic acid (imidazole propionate)

Imidazole propionate, a hub in our diabetes-associated modules, has been shown to impair insulin signaling through activation of the p38γ/p62/mTORC1 pathway and to inhibit metformin-induced AMPK activation, thereby reducing the hypoglycemic efficacy of metformin [57, 58]. Its association with the Bacteroides 2 enterotype and reduced gut bacterial gene richness implicates gut dysbiosis as an upstream driver of its accumulation [59], and polysaccharides from mulberry leaves that improve glycemia in diabetic rat models downregulate imidazole propionate among other metabolites [60]. Together with crotonic acid and glyoxylic acid, our data support a model in which gut microbiota-derived short-chain organic acids drive interconnected network hubs that impair insulin signaling and promote glucose dysregulation across the diabetes spectrum.

### Small peptide biology

Among additional novel findings, two micro-peptides, namely Ala-Asp-Gly-Gly (ADGG) and Leu-Arg-Thr-Gln-Val (LRTQV), displayed significant group-by-time interaction effects, with LRTQV showing a significant negative interaction selectively in the prediabetes group under our most stringent criteria. The structural features of these peptides suggest potential ligand activity: glycine’s flexibility facilitates induced-fit binding by small molecules [61], arginine and aspartate contribute to integrin-binding RGD-like motifs [62], and threonine hydroxyl groups enable receptor hydrogen bonding [63]. These micro-peptides represent potentially tractable biomarkers of early prediabetes and/or diabetes and warrant further investigation.

The main limitation of this study is the relatively small sample size (n=30 x 2 timepoints). However, the dynamic assessment of responses to a physiologically relevant glucose challenge, the deep phenotyping of the ORIGINS cohort, and the unparalleled depth of metabolomics profiling (>700 annotated metabolites consistently detected) substantially strengthen the biological relevance of the statistically significant findings reported here.

Altogether, this study identifies molecular networks underlying the development of prediabetes and type 2 diabetes, with short-chain organic acids (centered on crotonic acid) forming the pathophysiologically relevant network hubs. The progressive accumulation of crotonic acid, glyoxylic acid, 3S,4-dihydroxybutyric acid, imidazole propionate, and related organic acids, paired with deregulated long-chain fatty acid oxidation and altered amino acid metabolism, constitutes a composite metabolic signature of the diabetic state. The validation of this signature through its association with global protein crotonylation in circulating EV proteins establishes a mechanistic link between the serum metabolome and a broad post-translational modification program with demonstrated relevance to insulin signaling and glucose metabolism. Consequently, a unifying hypothesis develops: diabetes may involve coordinated subclinical deficiencies in the metabolism of glycine, serine, GABA, branched-chain amino acids, and short-chain fatty acids that collectively alter global protein crotonylation and impair glucose regulation. Future investigations in larger cohorts are needed to validate these hub metabolites as therapeutic targets, biomarkers of progression and/or response to treatment, and/or biomarkers for early detection in prediabetes and type 2 diabetes.

## Materials and Methods

### Sample Selection and Criteria

The AdventHealth Institutional Review Board approved all procedures. Prior to commencing the study, all volunteers provided informed consent. The study leveraged archived serum samples and richly annotated clinical data from 30 human subjects with 10 individuals per group from the ORIGINS (Uncovering the ORIGINS of Diabetes) study (ClinicalTrials.gov identifier: NCT02226640). ORIGINS is a deeply phenotyped observational cohort designed to elucidate molecular heterogeneity across the diabetes spectrum through integrated metabolic, clinical, and multiomic characterization. The groups were comprised of individuals with normal glucose tolerance (NGT), prediabetes (PDM), or type 2 diabetes (T2D), as defined by the American Diabetes Association [64]. Subjects with type 1 diabetes or any other form of diabetes were excluded as per the ORIGINS study criteria. The participants were selected to assure the groups were well-balanced for age, sex, and obesity and were chosen through a partially automated process that utilized a custom script based on the *MatchIt* package in the R programming environment. This same cohort was used for our previous EV proteomic and phosphoproteomic study [24].

### Clinical Measurements

Standardized protocols were followed to obtain the clinical and metabolic measurements of the study subjects [24]. Body composition was assessed by dual-energy x-ray absorptiometry (DXA, GE Lunar iDEXA, Madison, WI, USA). Fasting blood samples were collected, and a 75 g OGTT was conducted over a 2-hour period. On a separate visit, an insulin-modified frequently sampled intravenous glucose tolerance test (FSIVGTT) was carried out. Plasma glucose levels were measured using the glucose oxidase method on a YSI 2300 STAT Plus Analyzer (YSI Life Sciences, Yellow Springs, OH, USA). Plasma insulin and C-peptide concentrations were determined utilizing the MSD human insulin assay kit and C-peptide kit, respectively (MSD, Rockville, MD, USA). HbA1c levels were measured by means of a Cobas Integra 800 Analyzer (Roche, Basel, Switzerland). β-cell function was evaluated by computing HOMA-B, the insulinogenic index [ΔIns0–300/ΔGluc0–300], and the insulin and C-peptide areas under the curve (AUC) in response to OGTT. HOMA-IR was calculated as previously described to assess insulin activity [65, 66]. Data obtained from the FSIVGTT were utilized to determine insulin sensitivity (Si) and acute insulin response to glucose (AIRg) using the Minimal Model method by Bergman [67, 68].

### Metabolomics profiling

Plasma metabolites were extracted from 50 ul of plasma with 80% (1:1) MeOH:ACN containing ¹³C phenylalanine as an internal standard, and centrifuged at 10,000 × g for 30 min at 4 °C. The supernatants were dried and resuspended in 50 µL of 50% acetonitrile/water for analysis using a Q Exactive Plus Orbitrap MS (QE MS) equipped with an HESI II probe and coupled to an Ultra High Pressure Liquid Chromatography system (Dionex UltiMate 3000). Samples were analyzed within 24 hours of reconstitution. Four modes (2x2 matrix) were implemented: HILIC and Reverse Phase C18 columns, both with positive and negative electrospray ionization. For quality control (QC) of samples, the Standard Reference Material SRF1950 was used, and three pooled serum samples were prepared from 10 uL aliquots from all samples in each subgroup (n=10). Reverse-phase (RP) chromatography was performed using an HSS C18 column (150 3 2.1 mm i.d., 1.7 mm; Waters) with a flow rate of 0.4 mL/min. Solvent A was 0.1% formic acid in water, and solvent B was 0.1% ACN. The LC gradient included a 1-minute hold at 0.1% B, followed by a ramp from 0.1% to 50% B over the next 7 minutes, and then a ramp to 99% over the following 8 min. A 3 min hold at 99% was followed by a return to 0% over the next 0.5 min. The run was completed with a 7 min reconditioning at 0.1% B. The RP separation was performed over 22 min in positive ionization mode. The mass spectrometer was operated in positive and negative ESI ion mode in the scan range of m/z 70 to 1,050 with the resolution of 70,000 at m/z 200, automatic gain control (AGC) target at 1.3 x 10^6^, and maximum injection time of 50 ms. The spray voltage was set to 3.5 and 2.5 kV in positive and negative ion mode, respectively. The heated capillary was set at 200°C, the HESI probe was set at 350°C, and the S-lens RF level was set at 45. The gas settings for sheath, auxiliary, and sweep were 40, 10 and 1 unit, respectively. Peak areas (AUC) were used for comparative quantitation. RAW data files were processed with Xcalibur Quan software (Thermo Scientific) and Progenesis Qi (Waters) for filtering, alignment, feature detection, normalization, and annotation of the LC-MS signals.

Putative metabolite ID was based on accurate mass (error<10 ppm) as compared to Metlin & NIST databases. Area counts for each analyte were used for analysis. For statistical analysis of the metabolomic data, the means of the quadruplicate measurements (one from each LC-MS mode) were used.

### Weighted Coexpression Network Analysis

Weighted gene correlation network analysis, or WGCNA, is an algorithm and R package that identifies clusters of interrelated genes using graphs and networks composed of pairwise correlations of expression levels [25]. While WGCNA is primarily used for analyzing genomic data, we applied the algorithm towards metabolomic data. This scenario is not an uncommon use case, and prior research has shown WGCNA to be a suitable and unique method of analyzing metabolomic data [69]. Rather than focusing on individual metabolites, as in traditional differential abundance analyses, WGCNA takes advantage of the underlying correlations between metabolites and groups metabolites that show similar expression patterns and are directly or indirectly related to one another. These sets of related metabolites are then assigned to modules, which are correlated to clinical variables to determine trait significance and module membership, providing valuable clinical insight.

Following the procedure outlined by Almeida-Silva, et al. in the *BioNERO* R package (a streamlined library for WGCNA [26]) documentation, we constructed signed coexpression networks and identified modules for the Baseline metabolite measurement dataset (n = 30, 10 in each cohort) and the Delta metabolite measurement dataset (n=30, 10 in each cohort), which represents the difference in expression of metabolites at OGTT-120min and Baseline. The datasets all contained measurements of the 709 identifiable metabolites at their respective timepoints. This process resulted in two different coexpression networks, each with their own unique modules.

The module identification process began with inputting and cleaning the datasets (quantile normalization, outlier identification, etc.). Next, a soft power of β = 5 and 12 was calculated for the Baseline and Delta datasets on the basis of scale-free topology, respectively. The co-expression similarity was raised to this power to calculate adjacencies. Topological overlap matrices were then calculated from the adjacency matrices, allowing for both direct and indirect relationships between metabolites to be gathered. The deepSplit parameter was set to 2, and a minimum module size of 10 metabolites was used. Hierarchical clustering was employed to generate dendrograms displaying metabolite relationships, and this tree was subsequently cut (height = 0.25) to determine relevant metabolite modules. Lastly, the identified modules in Baseline and Delta were correlated with clinical variables. Furthermore, modules of metabolites with significant (p < 0.05) correlations to diabetes-relevant clinical traits (diabetes group, C-peptide, HbA1c, glucose levels, HOMA-IR, etc.) were selected for downstream analyses.

### Metabolite Set Enrichment Analysis

Metabolite set enrichment analysis (MSEA) was performed using the enrichment analysis R package MetaboAnalystR [27]. Specifically, MSEA was conducted on each of the significant modules identified in the baseline and delta datasets to determine biological pathway and mechanistic relevance. The list of 709 identified metabolites was used as the background metabolite set. The accepted descriptions for these metabolites were matched to their Small Molecule Pathway Database (SMPDB) IDs [28]. Correspondingly, the SMPDB metabolic pathways were chosen as the metabolite set library for matching.

### Extracellular Vesicle (EV) Proteomics and Protein Crotonylation Analysis

Our group previously reported mass spectrometry methods used for EV proteomic profiling of the same serum samples used in this metabolomic study [24]. Because crotonic acid was one of the key metabolites uncovered by this study (results to be presented later in this manuscript), we integrated the analysis of EV crotonylated proteins here. In brief, EVs were isolated using EVtrap® technology from Tymora Analytical Operations, and proteins were identified and quantified using an Ultimate 3000 nano UHPLC system (Thermo Fisher Scientific) coupled with a Q-Exactive HF-X mass spectrometer (Thermo Fisher Scientific). The mass spectrometer was operated in the data-dependent mode, and a full-scan MS (from m/z 375 to 1,500 with the resolution of 60,000) was followed by MS/MS of the 15 most intense ions with a resolution of 30,000. The raw files were searched directly against the human UniProt database with no redundant entries, using Byonic (Protein Metrics) and Sequest search engines loaded into Proteome Discoverer 2.3 software (Thermo Fisher Scientific). MS1 precursor mass tolerance was set at 10 ppm, and MS2 tolerance was set at 20 ppm. Search criteria to identify crotonylated proteins included a mass shift (+68.0375 Da) associated with the addition of a crotonyl group to lysine residues. The search was performed with full trypsin/P digestion and allowed a maximum of two missed cleavages on the peptides analyzed in the sequence database. The false discovery rates of proteins and peptides were set at 0.01. All protein and peptide identifications were grouped, and any redundant entries were removed. Only unique peptides and unique master proteins were reported.

### Statistical Analysis

Metadata normality was tested using the Shapiro-Wilk test, and non-normal data were log-transformed to approximate normality. Differences in baseline clinical characteristics were assessed using the Welch two-sample t-test (for continuous variables) or the Fisher exact test (for categorical variables). Analysis of differential abundance of metabolites and crotonylated proteins was conducted using linear models implemented with the *limma* package [70]. The linear models for the longitudinal OGTT metabolomics data included a group-by-time interaction term to account for the potential group-specific differential effects in response to the glucose challenge over time. To account for the correlation between the repeated measures at baseline and OGTT-120min, *duplicateCorrelation* was calculated and used to model the random effects of the Subject_ID variable. The design matrix was specified as:

design <- model.matrix(∼ DM_Group + DM_Group:time, metadata),

where DM_Group has 3 levels (NGT = reference, PreT2D, T2D) and time has 2 levels (“0” = reference, “120”). This is a reference-within-group interaction design. The key architectural choice is the absence of a main effect for time. Using ∼ DM_Group + DM_Group:time, instead of ∼ DM_Group * time, causes R to absorb the time effect entirely into the group:time interaction terms, giving each group its own independent temporal slope from baseline [71, 72]. This parameterization estimates group specific response trajectories rather than a pooled time effect, which is preferable for longitudinal designs such as OGTT, where temporal dynamics are expected to differ by group. The design columns interpretation is as follows:

| # | Column | Interpretation |
| --- | --- | --- |
| 1 | (Intercept) | Mean for NGT at time 0 (grand baseline) |
| 2 | DM_GroupPreT2D | PreT2D - NGT difference at time 0 (fasting) |
| 3 | DM_GroupT2D | T2D - NGT difference at time 0 (fasting) |
| 4 | DM_GroupNGT:time120 | Within-NGT change from time 0 to 120 (OGTT response) |
| 5 | DM_GroupPreT2D:time120 | Within-PreT2D change from time 0 to 120 |
| 6 | DM_GroupT2D:time120 | Within-T2D change from time 0 to 120 |

A full contrast matrix was then generated using the *makeContrasts* function of the *limma* package to assess the relevant comparisons:

| Contrast | Vector | Biological Question |
| --- | --- | --- |
| PreT2D_vs_NGT.t0 | [ 0, +1, 0, 0, 0, 0] | Fasting difference: PreT2D vs NGT |
| T2D_vs_NGT.t0 | [ 0, 0, +1, 0, 0, 0] | Fasting difference: T2D vs NGT |
| T2D_vs_PreT2D.t0 | [ 0, -1, +1, 0, 0, 0] | Fasting difference: T2D vs PreT2D |
| PreT2D_vs_NGT.t120 | [ 0, +1, 0, -1, +1, 0] | Post-OGTT: PreT2D vs NGT |
| T2D_vs_NGT.t120 | [ 0, 0, +1, -1, 0, +1] | Post-OGTT: T2D vs NGT |
| T2D_vs_PreT2D.t120 | [ 0, -1, +1, 0, -1, +1] | Post-OGTT: T2D vs PreT2D |
| Interaction.PreT2D_vs_NGT | [ 0, 0, 0, -1, +1, 0] | Is PreT2D OGTT response != NGT? |
| Interaction.T2D_vs_NGT | [ 0, 0, 0, -1, 0, +1] | Is T2D OGTT response != NGT? |
| Interaction.T2D_vs_PreT2D | [ 0, 0, 0, 0, -1, +1] | Is T2D OGTT response != PreT2D? |

The interaction terms are the highest-value contrasts as they test whether the molecular response to the glucose challenge differs across the glycemic spectrum (NGT, Prediabetes, T2D). The Benjamini-Hochberg correction, as implemented for the *p.adjust* function in the *stats* package, was used to calculate the false discovery rate (FDR) and correct for multiple testing.

Metabolomic features with fold change FC>1.5, P<0.05, and FDR<0.05 were considered significant. Correlations between serum metabolites and serum crotonylated EV proteins with absolute r>0.4, P<0.05, and FDR<0.01 were considered significant.

## Data availability

All data produced in the present study will be available upon reasonable request to the authors after actual publication in peer-reviewed journal.

## Author Contributions

REP and YNL designed and supervised the study. REP funded the study, interpreted results, and critically reviewed the manuscript. YNL and DD wrote the R scripts, analyzed data, interpreted results, and wrote the manuscript. TD conducted mass spectrometry experiments for metabolomics, generated and analyzed data, interpreted results, and critically reviewed the manuscript. AI conducted mass spectrometry experiments for EV proteomics, generated data, interpreted results, and critically reviewed the manuscript. AC analyzed data, interpreted results, contributed to manuscript writing, and critically reviewed the manuscript. SJG supervised the metabolomics experiments, interpreted results, and critically reviewed the manuscript.

## Acknowledgements

The authors thank Dr. Steven R. Smith for providing access to the archived samples from the ORIGINS study.

## References

1. Khan, M.A.B., et al., Epidemiology of Type 2 Diabetes - Global Burden of Disease and Forecasted Trends. J Epidemiol Glob Health, 2020. 10(1): p. 107–111.

2. Hostalek, U., Global epidemiology of prediabetes - present and future perspectives. Clin Diabetes Endocrinol, 2019. 5: p. 5.

3. Chatterjee, S., K. Khunti, and M.J. Davies, Type 2 diabetes. Lancet, 2017. 389(10085): p. 2239–2251.

4. Gilmer, T.P., et al., Predictors of health care costs in adults with diabetes. Diabetes Care, 2005. 28(1): p. 59–64.

5. Goyal, R., M. Singhal, and I. Jialal, Type 2 Diabetes, in StatPearls. 2025: Treasure Island (FL).

6. Vas, P.R.J., K.G. Alberti, and M.E. Edmonds, Prediabetes: moving away from a glucocentric definition. Lancet Diabetes Endocrinol, 2017. 5(11): p. 848–849.

7. Tabak, A.G., et al., Prediabetes: a high-risk state for diabetes development. Lancet, 2012. 379(9833): p. 2279–90.

8. Whiting, D.R., et al., IDF diabetes atlas: global estimates of the prevalence of diabetes for 2011 and 2030. Diabetes Res Clin Pract, 2011. 94(3): p. 311–21.

9. Passaro, A.P., et al., Omics era in type 2 diabetes: From childhood to adulthood. World J Diabetes, 2021. 12(12): p. 2027–2035.

10. Chen, R., et al., LC-MS-Based Untargeted Metabolomics Reveals Early Biomarkers in STZ-Induced Diabetic Rats With Cognitive Impairment. Front Endocrinol (Lausanne), 2021. 12: p. 665309.

11. Milburn, M.V. and K.A. Lawton, Application of metabolomics to diagnosis of insulin resistance. Annu Rev Med, 2013. 64: p. 291–305.

12. Klein, M.S. and J. Shearer, Metabolomics and Type 2 Diabetes: Translating Basic Research into Clinical Application. J Diabetes Res, 2016. 2016: p. 3898502.

13. Chen, Z.-Z. and R.E. Gerszten, Metabolomics and Proteomics in Type 2 Diabetes. Circulation Research, 2020. 126(11): p. 1613–1627.

14. Newgard, C.B., et al., A branched-chain amino acid-related metabolic signature that differentiates obese and lean humans and contributes to insulin resistance. Cell Metab, 2009. 9(4): p. 311–26.

15. Wang, T.J., et al., Metabolite profiles and the risk of developing diabetes. Nat Med, 2011. 17(4): p. 448–53.

16. Newgard, C.B., Interplay between lipids and branched-chain amino acids in development of insulin resistance. Cell Metab, 2012. 15(5): p. 606–14.

17. Lynch, C.J. and S.H. Adams, Branched-chain amino acids in metabolic signalling and insulin resistance. Nat Rev Endocrinol, 2014. 10(12): p. 723–36.

18. Merino, J., et al., Metabolomics insights into early type 2 diabetes pathogenesis and detection in individuals with normal fasting glucose. Diabetologia, 2018. 61(6): p. 1315–1324.

19. Li, J., et al., Circulating metabolites, genetics and lifestyle factors in relation to future risk of type 2 diabetes. Nat Med, 2026. 32(2): p. 660–670.

20. Sharma, S., et al., Role of human plasma metabolites in prediabetes and type 2 diabetes from the IMI-DIRECT study. Diabetologia, 2024. 67(12): p. 2804–2818.

21. Cui, L., H. Lu, and Y.H. Lee, Challenges and emergent solutions for LC-MS/MS based untargeted metabolomics in diseases. Mass Spectrom Rev, 2018. 37(6): p. 772–792.

22. Steuer, R., Review: on the analysis and interpretation of correlations in metabolomic data. Brief Bioinform, 2006. 7(2): p. 151–8.

23. Nunez Lopez, Y.O., G. Garufi, and A.A. Seyhan, Altered levels of circulating cytokines and microRNAs in lean and obese individuals with prediabetes and type 2 diabetes. Mol Biosyst, 2016. 13(1): p. 106–121.

24. Nunez Lopez, Y.O., et al., Proteomics and Phosphoproteomics of Circulating Extracellular Vesicles Provide New Insights into Diabetes Pathobiology. Int J Mol Sci, 2022. 23(10).

25. Langfelder, P. and S. Horvath, WGCNA: an R package for weighted correlation network analysis. BMC Bioinformatics, 2008. 9: p. 559.

26. Almeida-Silva, F. and T.M. Venancio, BioNERO: an all-in-one R/Bioconductor package for comprehensive and easy biological network reconstruction. Functional & Integrative Genomics, 2022. 22(1): p. 131–136.

27. Chong, J. and J. Xia, MetaboAnalystR: an R package for flexible and reproducible analysis of metabolomics data. Bioinformatics, 2018. 34(24): p. 4313–4314.

28. Jewison, T., et al., SMPDB 2.0: big improvements to the Small Molecule Pathway Database. Nucleic Acids Res, 2014. 42(Database issue): p. D478–84.

29. Felig, P., E. Marliss, and G.F. Cahill, Jr., Plasma amino acid levels and insulin secretion in obesity. N Engl J Med, 1969. 281(15): p. 811–6.

30. Würtz, P., et al., Branched-Chain and Aromatic Amino Acids Are Predictors of Insulin Resistance in Young Adults. Diabetes Care, 2013. 36(3): p. 648–655.

31. Floegel, A., et al., Identification of Serum Metabolites Associated With Risk of Type 2 Diabetes Using a Targeted Metabolomic Approach. Diabetes, 2013. 62(2): p. 639–648.

32. Wang-Sattler, R., et al., Novel biomarkers for pre-diabetes identified by metabolomics. Mol Syst Biol, 2012. 8: p. 615.

33. Lee, H.J., et al., 2-Aminoadipic acid (2-AAA) as a potential biomarker for insulin resistance in childhood obesity. Sci Rep, 2019. 9(1): p. 13610.

34. Cheng, S., et al., Metabolite profiling identifies pathways associated with metabolic risk in humans. Circulation, 2012. 125(18): p. 2222–31.

35. Wang, T.J., et al., 2-Aminoadipic acid is a biomarker for diabetes risk. J Clin Invest, 2013. 123(10): p. 4309–17.

36. Guasch-Ferré, M., et al., Metabolomics in Prediabetes and Diabetes: A Systematic Review and Meta-analysis. Diabetes Care, 2016. 39(5): p. 833–846.

37. Erdbrügger, P. and F. Fröhlich, The role of very long chain fatty acids in yeast physiology and human diseases. Biological Chemistry, 2021. 402(1): p. 25–38.

38. Lu, Q., et al., *The Overlooked Transformation Mechanisms of VLCFAs: Peroxisomal* β*-Oxidation*. Agriculture, 2022. 12(7): p. 947.

39. Yamada, K. and T. Taketani, Management and diagnosis of mitochondrial fatty acid oxidation disorders: focus on very-long-chain acyl-CoA dehydrogenase deficiency. J Hum Genet, 2019. 64(2): p. 73–85.

40. Tan, M., et al., Identification of 67 histone marks and histone lysine crotonylation as a new type of histone modification. Cell, 2011. 146(6): p. 1016–28.

41. Yang, P., et al., Crotonylation and disease: Current progress and future perspectives. Biomedicine & Pharmacotherapy, 2023. 165: p. 115108.

42. Liao, P., et al., Crotonylation at serine 46 impairs p53 activity. Biochemical and Biophysical Research Communications, 2020. 524(3): p. 730–735.

43. Liu, Y., et al., Non-Histone Lysine Crotonylation Is Involved in the Regulation of White Fat Browning. International Journal of Molecular Sciences, 2022. 23(21): p. 12733.

44. Quaye, E., et al., Energy expenditure due to gluconeogenesis in pathological conditions of insulin resistance. American Journal of Physiology-Endocrinology and Metabolism, 2021. 321(6): p. E795–E801.

45. Lillegard, K., J.A. Del Castillo, and H.J. Silver, Poorly controlled glycemia and worse beta cell function associate with higher resting and total energy expenditure in adults with obesity and type 2 diabetes: A doubly labeled water study. Clinical Nutrition, 2024. 43(3): p. 729–738.

46. Liao, W., et al., Persistent high glucose induced EPB41L4A-AS1 inhibits glucose uptake via GCN5 mediating crotonylation and acetylation of histones and non-histones. Clin Transl Med, 2022. 12(2): p. e699.

47. Shan, S., et al., Marine algae-derived oligosaccharide via protein crotonylation of key targeting for management of type 2 diabetes mellitus in the elderly. Pharmacological Research, 2024. 205: p. 107257.

48. Belostotsky, R., J.J. Pitt, and Y. Frishberg, Primary hyperoxaluria type III—a model for studying perturbations in glyoxylate metabolism. Journal of Molecular Medicine, 2012. 90(12): p. 1497–1504.

49. Schnedler, N., G. Burckhardt, and B.C. Burckhardt, Glyoxylate is a substrate of the sulfate-oxalate exchanger, sat-1, and increases its expression in HepG2 cells. Journal of Hepatology, 2011. 54(3): p. 513–520.

50. Nikiforova, V.J., et al., Glyoxylate, a new marker metabolite of type 2 diabetes. J Diabetes Res, 2014. 2014: p. 685204.

51. Song, S., Can the glyoxylate pathway contribute to fat-induced hepatic insulin resistance? Med Hypotheses, 2000. 54(5): p. 739–47.

52. Padberg, I., et al., A new metabolomic signature in type-2 diabetes mellitus and its pathophysiology. PLoS One, 2014. 9(1): p. e85082.

53. Franklin, I.K. and C.B. Wollheim, GABA in the endocrine pancreas: its putative role as an islet cell paracrine-signalling molecule. J Gen Physiol, 2004. 123(3): p. 185–90.

54. Michalik, M. and M. Erecińska, Gaba in pancreatic islets: Metabolism and function. Biochemical Pharmacology, 1992. 44(1): p. 1–9.

55. Curovic, V.R., et al., Circulating Metabolites and Lipids Are Associated to Diabetic Retinopathy in Individuals With Type 1 Diabetes. Diabetes, 2020. 69(10): p. 2217–2226.

56. Hou, D. and Y. Yang, Genetically predicted elevated circulating 3,4-dihydroxybutyrate levels mediate the association between family Christensenellaceae and osteoporosis risk: a Mendelian randomization study. Front Endocrinol (Lausanne), 2024. 15: p. 1388772.

57. Molinaro, A., et al., Imidazole propionate is increased in diabetes and associated with dietary patterns and altered microbial ecology. Nature Communications, 2020. 11(1): p. 5881.

58. Koh, A., et al., Microbially Produced Imidazole Propionate Impairs Insulin Signaling through mTORC1. Cell, 2018. 175(4): p. 947–961.e17.

59. Le Chatelier, E., et al., Richness of human gut microbiome correlates with metabolic markers. Nature, 2013. 500(7464): p. 541–546.

60. Ma, Q., et al., Therapeutic mechanisms of mulberry leaves in type 2 diabetes based on metabolomics. Front Pharmacol, 2022. 13: p. 954477.

61. Katsoulidis, A.P., et al., Chemical control of structure and guest uptake by a conformationally mobile porous material. Nature, 2019. 565(7738): p. 213–217.

62. D’Souza, S.E., M.H. Ginsberg, and E.F. Plow, Arginyl-glycyl-aspartic acid (RGD): a cell adhesion motif. Trends Biochem Sci, 1991. 16(7): p. 246–50.

63. Ippolito, J.A., R.S. Alexander, and D.W. Christianson, Hydrogen bond stereochemistry in protein structure and function. Journal of Molecular Biology, 1990. 215(3): p. 457–471.

64. American Diabetes Association Professional Practice, C., 2. Diagnosis and Classification of Diabetes: Standards of Care in Diabetes-2025. Diabetes Care, 2025. 48(1 Suppl 1): p. S27–S49.

65. Wallace, T.M., J.C. Levy, and D.R. Matthews, Use and abuse of HOMA modeling. Diabetes Care, 2004. 27(6): p. 1487–95.

66. Prystupa, K., et al., Comprehensive validation of fasting-based and oral glucose tolerance test-based indices of insulin secretion against gold standard measures. BMJ Open Diabetes Res Care, 2022. 10(5).

67. Pacini, G., et al., Insulin sensitivity and glucose effectiveness: minimal model analysis of regular and insulin-modified FSIGT. American Journal of Physiology-Endocrinology and Metabolism, 1998. 274(4): p. E592–E599.

68. Bergman, R.N., et al., Quantitative estimation of insulin sensitivity. Am J Physiol, 1979. 236(6): p. E667–77.

69. Pei, G., L. Chen, and W. Zhang, WGCNA Application to Proteomic and Metabolomic Data Analysis. Methods Enzymol, 2017. 585: p. 135–158.

70. Ritchie, M.E., et al., *limma powers differential expression analyses for RNA-sequencing and microarray studies*. Nucleic Acids Research, 2015. 43(7): p. e47–e47.

71. Law, C.W., et al., A guide to creating design matrices for gene expression experiments. F1000Res, 2020. 9: p. 1444.

72. Smyth, G.K., Linear models and empirical bayes methods for assessing differential expression in microarray experiments. Stat Appl Genet Mol Biol, 2004. 3: p. Article3.

